# Association of thyroid autoimmunity and the response to recombinant human growth hormone in Turner syndrome

**DOI:** 10.1101/2020.09.13.20193573

**Authors:** Yuyao Song, Hongbo Yang, Linjie Wang, Fengying Gong, Hui Pan, Huijuan Zhu

**Author notes:** Corresponding Author: Huijuan Zhu Department of Endocrinology, Peking Union Medical College Hospital, Shuaifuyuan No. 1, Dongcheng district, Beijing, 100005, China.

## Abstract

**Introduction:** Short stature and thyroid autoimmunity are among the most common traits in Turner syndrome (TS). Recombinant human growth hormone (rhGH) treatment benefits height growth in Turner syndrome individuals when applicable. This study aims to investigate the association of thyroid autoimmunity and the response to rhGH treatment in Turner Syndrome patients.

**Methods:** Medical records of 494 patients with TS were reviewed. Among 126 patients who regularly tested for thyroid autoantibodies, 108 patients had received rhGH treatment. Clinical characteristics, including karyotype and the presence of autoimmune thyroid diseases, as well as rhGH treatment records were analyzed. Height velocity (HV) of patients with or without thyroid autoimmunity was compared to assess the response to rhGH treatment. For patients who received rhGH treatment and positive for thyroid autoantibodies, height velocity before and after antibody presence was compared.

**Results:** 45XO monosomy presented in 36% (176/496) of patients. 42.1% of patients (53/126) had elevated circulating anti-thyroid peroxidase antibody (TPOAb) and anti-thyroglobulin antibody (TgAb). In 108 patients who received rhGH treatment, a negative correlation was found between circulating TPOAb concentration and HV (n=53, r = -0.276, P<0.05). For patients who developed thyroid autoantibodies during rhGH treatment, HVs after thyroid autoantibody presence significantly decreased compared with HVs before thyroid autoantibody detection (n=44, p=0.0017).

**Conclusions:** Our data suggested that in preadult TS patients who developed thyroid autoantibodies during rhGH treatment, the response to rhGH is negatively associated with the development of thyroid autoimmunity.

## Introduction

Turner syndrome (TS) is one of the most common chromosomal diseases caused by lack of one X chromosome or structurally abnormal X chromosome. The frequency of TS is estimated as 25-50 per 100,000 females [1]. Clinical manifestations include growth retardation, characteristic appearances, delayed puberty, incomplete sexual developments and infertility [1].

Short stature is one of the main challenges in the clinical management of Turner syndrome. The spontaneous growth of most TS patients lagged far behind normal females. Short stature of TS patients was thought to be resulted from skeletal developmental defects [2] and could be treated by recombinant human growth hormone (rhGH) [1]. Early initiation of recombinant human growth hormone (rhGH) treatment is suggested to improve final height in TS patients [1, 3-5]. It was reported that rhGH treatment could improve the average height velocity (HV) from 2.9 cm/yr to 7.3 cm/yr in TS patients [3] and give an average final height gain of 7.2cm [1, 6]. There are several factors associated with rhGH treatment outcome, including the age at treatment start, duration of treatment, rhGH intake frequency and dosage [1, 3-5], and other genetic-associated factors such as mid-parental height (MPH) [1]. Also, delayed bone age was reported to be associated with better rhGH treatment outcome [5].

TS patients have increased risk of autoimmune disorders, such as inflammatory bowel disease, juvenile rheumatoid arthritis, type 1 diabetes mellitus and especially, autoimmune thyroid diseases (AITDs) [1]. TS patients are more likely to develop AITDs compared with normal female throughout childhood and adulthood [7, 8]. The overall prevalence of AITDs in TS patients is 38.6% and Hashimoto’s thyroiditis (HT) is the most common subtype [9].

HT patients automatically develop anti-thyroid peroxidase antibody (TPOAb) and anti-thyroglobulin antibody (TgAb) [9]. It was reported that TPOAb plays a main role for cell-mediated cytotoxicity associated with thyroid autoimmunity, leading to diffuse lymphatic infiltration, inflammation and dysfunction of the thyroid gland [10].

IGF-1 level is one of the major factors correlated with growth rate during rhGH treatment [11]. An increasing correlation between autoimmune diseases and IGF-1 resistance had been found recently [12]. The disruption of IGF-1 signaling pathway was reported in several autoimmune diseases [12-14]. Antibodies against IGF-1 receptors (IGF-1R) were detected in Graves’ disease [12]. In patients with type 1 diabetes, tissue-specific mutation of IGF-1R was found in pancreatic β cells [13]. In patients with rheumatic arthritis, IGF-1 or IgG collected from patients could activate disease-derived synovial fibroblasts and CD4+ T cells suggested that the peripheral tolerance of IGF-1R was broken [14]. Although the IGF-1 signaling pathway disruption plays a role in autoimmunity, the association of thyroid autoimmunity and the response to rhGH treatment in Turner syndrome has not been investigated yet.

In this single-center retrospective study, we analyzed the clinical characteristics and the height velocity of TS patients with different thyroid autoimmunity status during rhGH treatment.

## Materials and Methods

### Subjects

A retrospective chart review was performed in 496 consecutive patients with Turner syndrome (TS) followed up from 1999 to 2019 in Peking Union Medical College Hospital [15]. The diagnosis of TS was confirmed by karyotype with 45XO, or other structural abnormalities of the X chromosome. Among 126 patients who regularly tested TPOAb and TgAb every 3-6 months, 108 had received rhGH treatment. Approval from the Institutional Review Board of Peking Union Medical College Hospital was obtained for this study. All data were de-identified before analysis.

### Anthropometrics

Height and weight were measured by standard protocols in the early morning. BMI was calculated as weight (kg) divided by height (m) squared. Height velocity (HV, cm/yr) was calculated by average height growth (cm) per year to evaluate the response to rhGH treatment [16, 17].

### Biochemical measurements

Complete blood count, serum lipid profile, fasting blood glucose, uric acid, hepatic function and renal function tests were all done in the central laboratory of the PUMCH by standard methods. Serum concentration of free triiodothyronine (FT3), free thyroxine (FT4), thyroid-stimulating hormone (TSH), TPOAb and TgAb were measured by the ADVIA Centaur XP Immunoassay system (Siemens, Germany). Serum IGF-1 level was measured with a chemiluminescent enzyme immunometric assay (Immulite 2000, Siemens Healthcare Diagnostics, UK). All assays were performed in the department of clinical laboratory of PUMCH.

### Statistical analysis

Continuous data were represented by mean ± standard deviation (SD). Data analyses were performed with Microsoft Excel, version for windows (Microsoft Co. Ltd., Seattle, WA, USA) and SPSS 24.0 software, version for Windows (SPSS Inc., Chicago, IL, USA). Correlation of two variables was assessed with Pearson correlation coefficient (r). Differences between two groups were detected using independent sample, two-sided student’s t-test. Results with P<0.05 were considered statistically significant.

## Results

### Demographic, clinical and biochemical characteristics

As shown in Figure 1, medical charts of a total of 494 cases of TS patients were reviewed in our center. Among them, 36% (176/494) had 45XO monosomy, 32% (158/494) was chimeras, 10% (48/494) had X isochromosome, 13% (67/494) had partial deletion of X chromosome and 9% (45/494) had other X chromosome structural abnormalities (Table 1).

**Figure 1.**
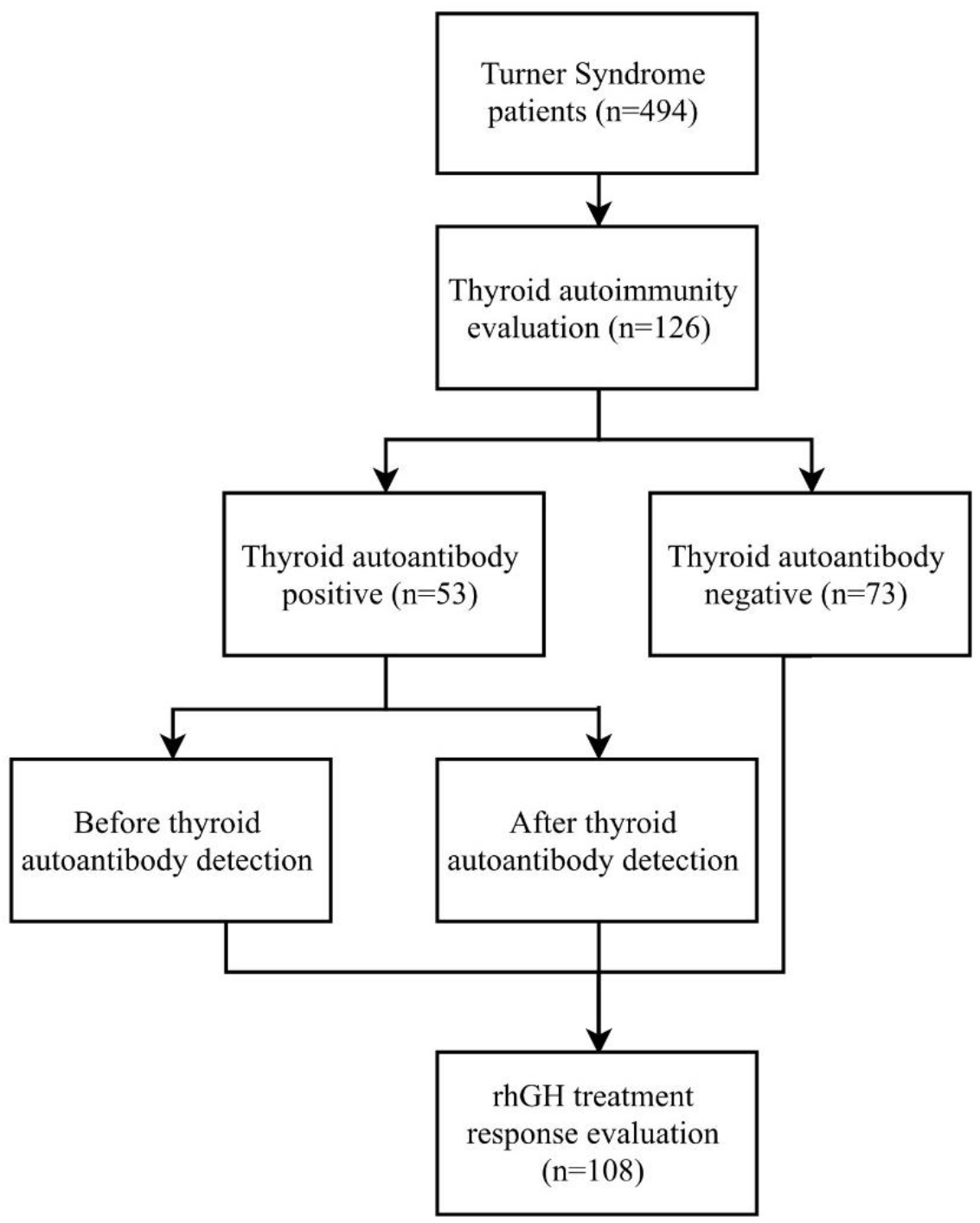
The flowchart of this study.

**Table 1.** Frequency of karyotypes of 494 TS patients in PUMCH

| Classification | Karyotype | Number | Frequency |
| --- | --- | --- | --- |
| X chromosome monosomy | 45XO | 176 | 36% |
| Chimeras | 45XO, 46XX | 50 | 32% |
|  | 45XO, 46X, i(Xq) | 57 |  |
|  | Other chimeras | 51 |  |
| X Isochromosome | 46X, Xi | 48 | 10% |
| Partial deletion of X chromosomes | 46X, del (Xq) | 32 | 13% |
|  | Mar | 35 |  |
| Others | Other | 45 | 9% |
| Total |  | 494 | 100% |

TS, turner syndrome; PUMCH, Peking Union Medical College Hospital

Among these patients, 126 cases had repeated thyroid autoantibodies test every three to six months during follow up and the general characteristics of these patients were summarized in Table 2. The age of diagnosis of TS was 10.4±4.1yr in this population. 42.1% of patients (53/126) had elevated TPOAb and TgAb and the first appearance of thyroid autoantibodies were around 12.6±4.9yr. 57.9% of patients (73/126) were negative for thyroid autoantibodies. Most patients developed TPOAb and TgAb simultaneously, and only one patient was only TPOAb positive during follow-up. The TPOAb concentrations varied widely (0-5000U/ml) among patients. Based on the presence of thyroid autoantibodies, patients were divided into two groups. There was no difference in the initial demographic, clinical and biochemical characteristics between the two groups of patients with or without thyroid autoantibodies (Table 2).

**Table 2.** Clinical and biochemical characteristics of 126 patients who regularly repeated thyroid autoantibodies tests

| Parameters | Thyroid auto-Ab positive<br>(n=53) | Thyroid auto-Ab negative<br>(n=73) | P value |
| --- | --- | --- | --- |
| Age (years) | 10.6±4.6 | 10.2±3.7 | 0.53 |
| Height (cm) | 124.8±19.3 | 124.3±14.6 | 0.60 |
| Weight (Kg) | 32.3±13.4 | 30.4±8.6 | 0.56 |
| BMI (kg/m<sup>2</sup>) | 19.6±3.7 | 19.0±2.7 | 0.38 |
| FT3 (pg/ml) | 3.72±0.56 | 3.85±0.76 | 0.29 |
| FT4 (ng/dl) | 1.30±0.18 | 1.71±2.08 | 0.13 |
| TSH (μIU/ml) | 3.84±4.48 | 3.37±3.49 | 0.54 |
| Hypothyroidism<sup>1</sup><br>(%) | 5.67 | 8.22 | 0.58 |
| Hyperthyroidism<br>(%) | 1.89 | 1.37 | 0.82 |
| TPOAb (IU/ml) | 113.09±202.72 | undetected | <0.01*** |
| TgAb (IU/ml) | 414.12±969.79 | undetected | <0.01*** |
| IGF-1 (ng/ml) | 439.65±189.71 | 483.35±213.75 | 0.32 |
| RBC (×10<sup>9</sup>/L) | 4.77±0.39 | 4.76±0.41 | 0.89 |
| WBC (×10<sup>9</sup>/L) | 6.62±1.87 | 6.52±1.96 | 0.80 |
| HGB (g/L) | 138.9±9.2 | 138.1±10.7 | 0.90 |
| PLT (×10<sup>9</sup>/L) | 291.10±78.27 | 284.51±82.86 | 0.68 |
| Cr (μmol/L) | 48.2±9.2 | 47.6±9.9 | 0.74 |
| Uric Acid (μmol/L) | 321.2±79.2 | 325.8±75.4 | 0.77 |
| ALT (U/L) | 19.4±14.7 | 19.6±12.7 | 0.94 |
| AST (U/L) | 25.7±9.1 | 27.8±7.6 | 0.33 |
| TBiL (μmol/L) | 10.1±4.9 | 10.8±8.4 | 0.58 |
| Glu (mmol/L) | 4.82±0.53 | 4.94±0.42 | 0.21 |
| Na (mmol/L) | 139.6±1.7 | 139.9±1.7 | 0.37 |
| K (mmol/L) | 4.3±0.3 | 4.4±0.3 | 0.31 |
| Ca (mmol/L) | 2.44±0.10 | 2.45±0.09 | 0.39 |
| P (mmol/L) | 1.60±0.22 | 1.73±0.20 | 0.005*** |
| ALP (U/L) | 203.5±91.2 | 230.0±80.1 | 0.24 |
| TC (mmol/L) | 4.60±0.72 | 4.75±0.85 | 0.58 |
| TG (mmol/L) | 1.02±0.66 | 1.05±0.70 | 0.90 |
| HDL-C (mmol/L) | 1.44±0.33 | 1.15±1.60 | 0.19 |
| LDL-C (mmol/L) | 2.60±0.57 | 1.96±2.58 | 0.94 |
| FSH (IU/L) | 55.59±39.00 | 69.00±45.02 | 0.19 |
| LH (IU/L) | 12.06±9.85 | 14.80±11.15 | 0.29 |
| E2 (IU/L) | 23.29±20.52 | 24.76±16.35 | 0.75 |

Data are represented as mean ± SD (\*\*: P<0.05, \*\*\*: P<0.01).
BMI, body mass index; rhGH, recombinant human growth hormone; FT3, free triiodothyronine; FT4, free thyroxine; TSH, thyroid stimulating hormone; TPOAb, thyroid peroxidase antibody; TgAb, anti-thyroglobulin antibody; IGF-1, insulin-like growth factor 1; RBC, red blood cell; WBC, white blood cell, HGB, hemoglobin, PLT, platelet; Cr, creatine; ALT, alanine aminotransferase; AST, aspartate aminotransferase; TBiL, total bilirubin; ALP, alkaline phosphatase; TC, total cholesterol; TG, triglyceride; HDL-C, high-density lipoprotein cholesterol; LDL-C, low-density lipoprotein cholesterol; FSH, follicle-stimulating hormone; LH, luteinizing hormone; E2, estrogen.

Among 126 patients, 84.1% (106/126) had a height lagged below the third percentile of their healthy peers. 85.7% (108/126) had received rhGH treatment, starting from 10.3±3.3 years old. The daily rhGH dosage was started from subcutaneous injection of 0.15 u/kg and titrated according to serum IGF-1 concentration and growth velocity. 52.8% (57/108) patients started rhGH treatment before 10 years old. In 53 patients who presented positive thyroid autoantibodies, 44 accepted rhGH treatment from 10.1±3.6 years old. While in 73 patients who were negative for thyroid autoantibodies, 64 accepted rhGH treatment from 10.5±3.2 years old in our clinic.

## Overall HV correlates to age, height, weight and BMI at treatment start point

For patients who received rhGH treatment (n=108), the treatment start age, rhGH dosage and the overall HV were not different between patients who developed thyroid autoimmunity and who did not (Table 3).

**Table 3.** Height velocity compared in rhGH treated TS patients with or without thyroid autoantibody

|  | Thyroid<br>auto-Ab positive | Thyroid<br>auto-Ab negative | P value |
| --- | --- | --- | --- |
| Number of subjects | 44 | 64 |  |
| Treatment start age<br>(yr) | 10.1±3.6 | 10.5±3.2 | 0.89 |
| rhGH treatment dose<br>(IU/kg/d) | 0.18 ±0.03 | 0.17± 0.04 | 0.59 |
| Height velocity<br>(cm/yr) | 6.53±2.56 | 6.12±2.27 | 0.39 |

Data are represented as mean±SD (\*\*: P<0.05, \*\*\*: P<0.01); rhGH, recombinant human growth hormone; TS, turner syndrome.

Considering treatment start age, patients who started rhGH treatment before 10 years old had better HV than patients who started rhGH treatment after 10 years old (7.55±1.66 vs 5.21±2.36 cm/yr, P<0.0001). A significant negative correlation between treatment start age and height velocity was also detected (n=108, rs=-0.65, P<0.01). For rhGH treated patients, significant negative correlations were found between HV and treatment start height (n=108, rs=-0.57, P<0.01), start weight (n=108, rs=-0.46, P<0.01) and start BMI (n=108, rs=-0.25, P<0.05).

## Thyroid autoimmunity status is associated with rhGH treatment response

However, for patients who developed thyroid autoantibodies during rhGH treatment, HVs after thyroid autoantibody presence significantly decreased compared with HVs before thyroid autoantibody presence (7.51±2.48 vs 5.51 ±2.39 cm/yr, P=0.0017, Figure 2 and Table 4). For rhGH treated thyroid auto-Ab positive patients, HV is significantly negatively correlated to treatment start age (n=44, rs=0.65, P<0.01), start height (n=44, rs=-0.62, P<0.01), start weight (n=44, rs=-0.54, P<0.01) and start BMI (n=44, rs=-0.38, P<0.05). A weak correlation was discovered between HV and serum TPOAb concentration for the same group of patients (n=44, rs = -0.22, P=0.15).

**Table 4.**
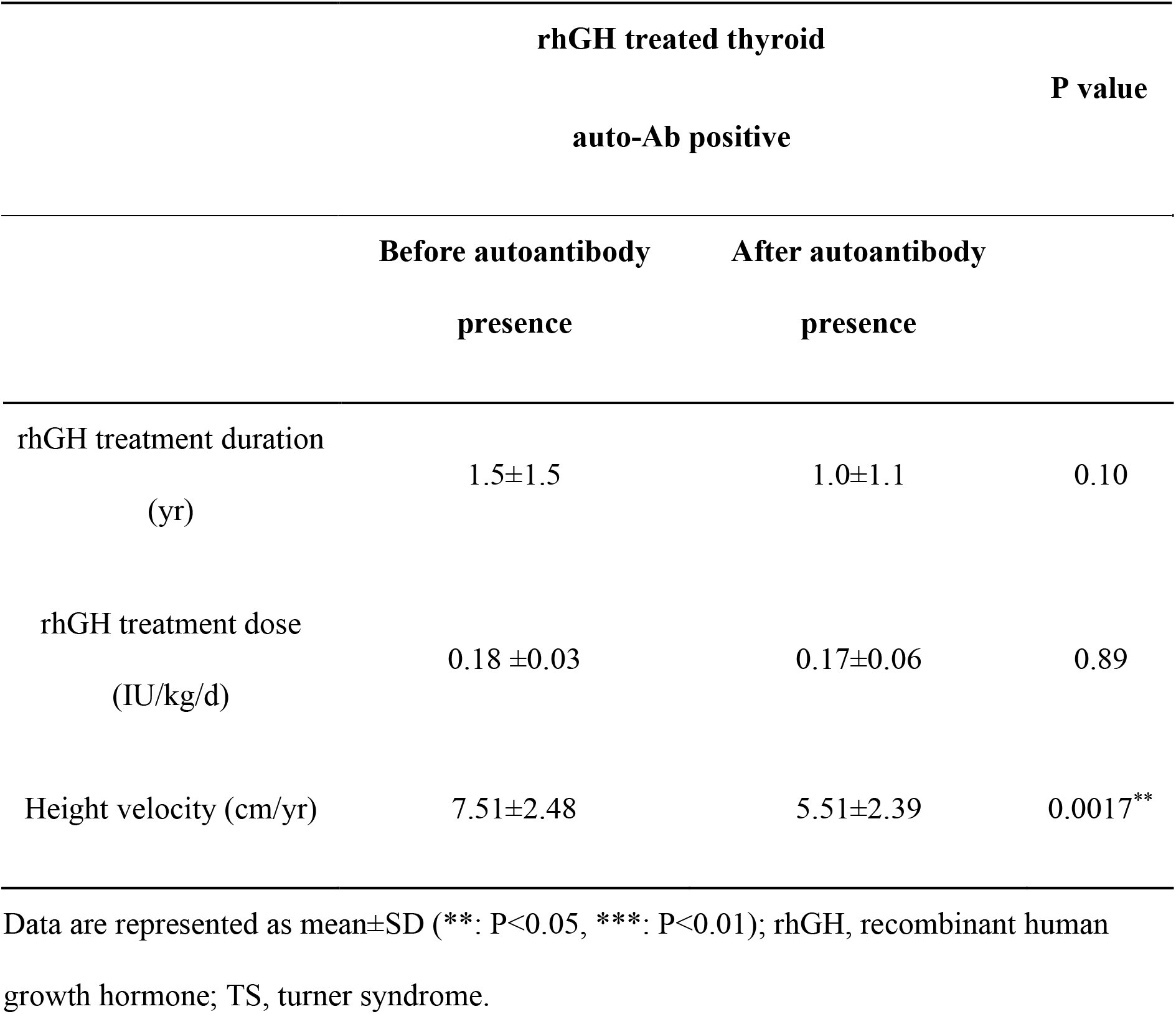
Height velocity compared in rhGH treated TS patients before and after presence of circulating thyroid autoantibodies

**Figure 2.**
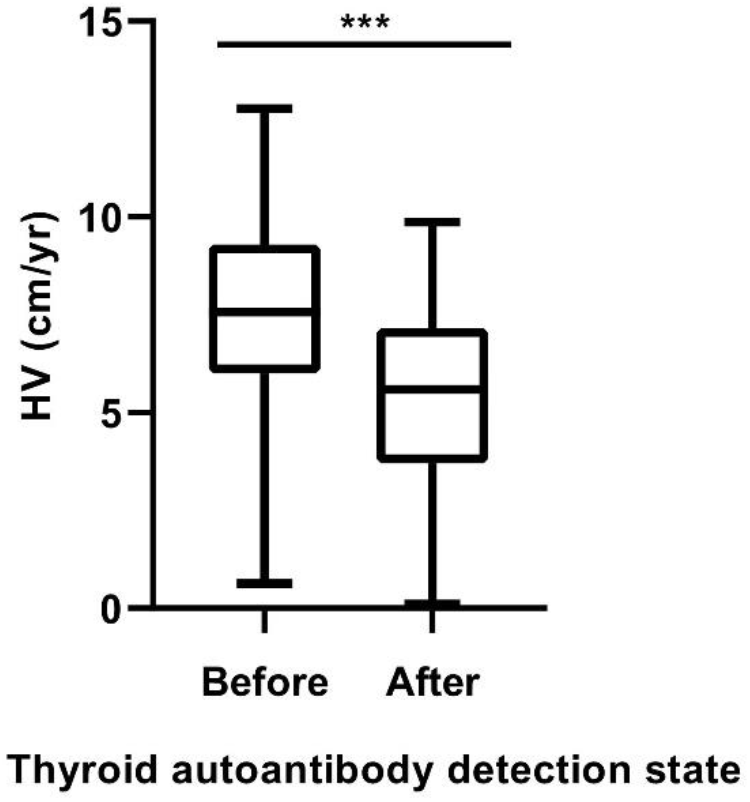
Height velocity (HV) in rhGH treated thyroid autoantibody positive TS patients before and after the detection of thyroid autoantibody. Patients have significantly lower HV after developing thyroid autoantibodies. Boxes indicate the first quartile and third quartile, whiskers indicated maximum and minimum values, means were represented by bars inside the boxes (n.s., not significant; **, P<0.05; ***, P<0.01).

## Discussion

In this retrospective single-center study, the clinical characteristics including karyotype and presence of autoimmune thyroid disease, as well as rhGH treatment records were analyzed in a cohort of patients with Turner syndrome. Both the distribution of karyotype [1, 18] and the average incidence of thyroid autoimmunity [8, 9, 18-20] are in consistence with previous studies. Our data further suggested that for TS patients who developed thyroid autoantibodies during rhGH treatment, the response to rhGH is negatively associated with the development of thyroid autoantibodies.

TS patients have significant high risk of autoimmune thyroid diseases [1, 7-9, 18-20]. TS children develop thyroid autoantibodies in a considerable high frequency and have higher prevalence of AITDs compared with the general pediatric population [7-9, 19, 20]. The morbidity of AITDs was found to increase significantly with age in TS patients and are absent among very young patients [7-9, 19, 20]. However, elevation of peripheral blood thyroid autoantibodies such as TPOAb and TgAb was always found at younger ages and priory to clinical symptoms of AITDs in TS patients [7, 8]. Our study is consistent with previous reports, with 42.06% of patients had elevated TPOAb and TgAb and the first appearance of thyroid autoantibodies around 12.6±4.9yr. The abnormal X chromosome composition has long been considered responsible for the increased risk of autoimmune disorders in TS patients [1]. But results of clinical studies of karyotype and thyroid autoimmunity are inconsistent [9, 18], and the underlying mechanism stayed unknown.

Plenty studies have discussed the factors associated with rhGH treatment outcome in TS patients. It was suggested that patients with higher mid-parental height, younger bone age, earlier treatment initiation, longer treatment duration before puberty induction and higher rhGH dosage have better outcomes in final height [1]. These have also been demonstrated in our data. Combination of ultra-low dose estrogen with rhGH had moderate benefit for height growth [21]. Meanwhile, the correlation of karyotype to rhGH treatment outcome remained obscure [22, 23] and the association of thyroid autoimmunity and the response to rhGH in TS patients is under-investigated so far. In this study, the average rhGH treatment start age was 10.31 years old. Previous reviews also found that in the general TS population, the rhGH treatment start age usually falls around 11.5 years old [9].

Our study firstly revealed that after presence of thyroid autoantibodies, height velocity decreased significantly in TS patients during rhGH treatment. The interplay of autoimmunity and childhood growth has been investigated from several aspects including the functional crosstalk between immune system and IGF-1 signaling pathway [24-27], and energy tradeoff between organ immune activity and growth [28-31].

The functional crosstalk of IGF-1 and immune regulation provides evidence from the aspect of signaling transduction. The IGF-1 signaling pathway acts through IGF-1, IGF-1 receptor (IGF-1R) and IGF binding proteins (IGFBPs) [24]. Relationship between immunity and IGF-1 signaling was detailly reviewed [24]. The review suggested that IGF-1 signaling pathway participate in the pathogenesis of autoimmune diseases in a complex and ambivalent manner [24]. Autoimmunity usually lead to the disruption of IGF-1/IGF-1R signaling, especially by producing autoantibodies against IGF-1R along with tissue-specific autoimmune targets [13, 24]. It was reported that in patients with GD [12, 14], T1DM [13] and RA [14], IGF-1R autoantibody development, IGF-1 resistance and tissue overexpression of IGF-1R were frequently observed as signals for the breakdown of peripheral tolerance of IGF-1R. Meanwhile, it was recently reported that immune derived cytokine signals might directly regulate certain IGF binding proteins (IGFBP), and immediately influence IGF-1 signaling [25]. Thus, for TS patients who developed thyroid autoimmunity, tissue might encounter IGF-1 resistance and have decreased response to rhGH treatment.

From the aspect of energy allocation, tissue immune activity is very energy costly and might derive energy from other activities [29]. Especially, physical growth was found most likely to be affected by the energy cost in tissue immunity during childhood [30-32]. A study had revealed that increased tissue activity in immune functions at childhood would impair height growth within a group of children living under high-pathogen, low-resource environments [31]. Since developing autoimmunity suggested hyperactive of immune system, height growth after thyroid autoimmunity might decrease compared with that of before in TS patients.

Recently, IGF-1 was found inhibitory of autoimmune diseases progression and inflammation through binding IGF-1R on immune cells [12, 26, 33]. However, the protective role of IGF-1 signaling was only found in limited circumstances [12]. It was reported that recombinant human insulin-like growth factor 1 (rhIGF-1) stimulates human and mouse regulatory T cells (Tregs) in vitro, and the delivery of rhIGF-1 to mouse models with type-1 diabetes and multiple sclerosis inhibits disease progression [26]. IGF-1 primed intestinal monocytes acquired immune inflammation suppressive ability by secreting IL-10 [33]. The reciprocal inhibition between autoimmunity and IGF-1 signaling pathway emphasized the importance of balance in the immune system.

Considering the above clinical and experimental results, alterations in treatment strategy should be made when supplementing rhGH to TS patients with autoimmune problems. However, more detailed clinical studies should be conducted to make suggestions. The decreased response might suggest increased treatment dose, but the IGF-1 level in TS patients should not be excessive [1]. The increasing risk of glucose intolerance and diabetes in TS patients alerted that rhGH treatment should be combined with a surveillance of IGF-1 level and glucose metabolism [1]. Combine with the fact that IGF-1 resistance is common in autoimmune diseases, we emphasize the importance of such surveillance epically in TS patients with thyroid autoimmunity.

The main limitation of this study is that our data did not provide a causal link between thyroid autoimmunity and response to rhGH treatment in TS patients since this is a retrospective study. Prospective studies are needed to further reveal the cause effect of thyroid autoimmunity in height growth among TS patients. At the same time, further studies to investigate the molecular mechanisms underlying relationship between Turner syndrome, thyroid autoimmunity and height growth are needed.

In summary, this study revealed a negative impact of thyroid autoimmunity on rhGH induced height growth on TS patients. Meanwhile, we also emphasized that an early treatment of rhGH significantly benefits patient height growth.

## Data Availability

The data that support the findings of this study are available from the corresponding author upon reasonable request. Due to ethical and privacy concerns, the data are not publicly available.

## Acknowledgement

The authors appreciate the CAMS Innovation Fund for Medical Science and the Non-profit Central Research Institute Fund of Chinese Academy of Medical Sciences for funding this research. We thank Dr. Yang Chen, Qi Wang and Zhenyi Wang for their visions and supports.

## Statement of Ethics

Written informed consents were given by parents and/or guardians of the studied subjects.

The study protocol was approved by the Tsinghua University School of Life Sciences committee on human research, and the Peking Union Medical College Hospital committee on human research.

## Author Contributions

Song YY carried out the investigation, performed data analysis, interpreted results and prepared drafts of the manuscript. Yang HB provided valuable insights in study design, helped data collection and made substantial contributions in manuscript preparation. Wang LJ, Gong FY and Pan H contributed to data acquisition and provided critical feedbacks. Zhu HJ contributed to study design and reviewed the manuscript.

